# Neuropsychiatric symptoms are associated with exacerbated cognitive impairment in covert cerebral small vessel disease

**DOI:** 10.1101/2021.12.20.21268088

**Authors:** Arola Anne, Levänen Tuuli, M. Laakso Hanna, Pitkänen Johanna, Koikkalainen Juha, Lötjönen Jyrki, Korvenoja Antti, Erkinjuntti Timo, Melkas Susanna, Jokinen Hanna

**Author notes:** Correspondence: Anne Arola, MSc, Department of Psychology and Logopedics, Faculty of Medicine, University of Helsinki, Finland.

## Abstract

**Background and purpose:** Neuropsychiatric symptoms are related to disease progression and cognitive decline over time in cerebral small vessel disease (SVD) but their significance is poorly understood in covert SVD. We investigated neuropsychiatric symptoms and their relationships between cognitive and functional abilities in subjects with varying degrees of white matter hyperintensities (WMH), but without clinical diagnosis of stroke, dementia or significant disability.

**Methods:** The Helsinki Small Vessel Disease Study consisted of 152 subjects, who underwent brain magnetic resonance imaging (MRI) and comprehensive neuropsychological evaluation of global cognition, processing speed, executive functions and memory. Neuropsychiatric symptoms were evaluated with the Neuropsychiatric Inventory Questionnaire (NPI-Q, n=134) and functional abilities with the Amsterdam Instrumental Activities of Daily Living questionnaire (A-IADL, n=132), both filled in by a close informant.

**Results:** NPI-Q total score correlated significantly with WMH volume (r_s_=0.20, p=0.019) and inversely with A-IADL score (r_s_=-0.41, p<0.001). In total, 38% of the subjects had one or more informant evaluated neuropsychiatric symptoms. Linear regressions adjusted for age, sex and education revealed no direct associations between neuropsychiatric symptoms and cognitive performance. However, there were significant synergistic interactions between neuropsychiatric symptoms and WMH volume on cognitive outcomes. Neuropsychiatric symptoms were also associated with A-IADL score irrespective of WMH volume.

**Conclusions:** Neuropsychiatric symptoms are associated with an accelerated relationship between WMH and cognitive impairment. Furthermore, the presence of neuropsychiatric symptoms is related to worse functional abilities. Neuropsychiatric symptoms should be routinely assessed in covert SVD as they are related to worse cognitive and functional outcomes.

## INTRODUCTION

Neuropsychiatric symptoms are highly prevalent in cognitive decline and dementia.^1,2^ Since neuropsychiatric symptoms are associated with higher risk of institutionalisation and even death, they are an extremely important factor to consider in clinical settings.^3^

Both cognitive decline and neuropsychiatric symptoms occur in cerebral small vessel disease (SVD). The consistent associations between SVD brain pathology (white matter hyperintensities [WMH], lacunes, cerebral microbleeds and brain atrophy) and cognitive impairment have been established in previous studies (for a recent review see ^4^). More severe WMH have been linked to apathy, fatigue and delirium as well as overall severity of neuropsychiatric symptoms.^5,6^ Apathy has been found to relate to SVD both in terms of disrupted white matter track connectivity and worse cognitive functioning.^7,8^ Overall SVD-related brain changes have been shown to relate to depression, and an association between white matter tract damage and depression has been found in patients with symptomatic SVD (including stroke).^9,10^

Previous research has shown that neuropsychiatric symptoms are associated with cognitive impairment. In community-dwelling subjects, cognitive impairment was related to higher incidence of neuropsychiatric symptoms and specific cognitive domains were associated with specific neuropsychiatric symptoms (executive functions and visuomotor speed with hyperactivity and affective symptoms; language and visual memory with psychosis symptoms).^11^ In overt SVD with dementia, higher prevalence of neuropsychiatric symptoms has been found to relate to worse cognitive outcomes.^12^

In a memory clinic sample of elderly subjects, who were free from dementia at baseline, increased WMH over time was associated with more neuropsychiatric symptoms, and an increase in both WMH and neuropsychiatric symptoms was related to cognitive decline over a two year period.^13^ Pathological brain changes such as WMH and lacunes have been associated with higher incidence of neuropsychiatric symptoms in patients with subcortical vascular impairment.^6^ In particular, WMH were found to relate to higher incidence of apathy and higher overall frequency of neuropsychiatric symptoms.

The majority of the previous research has focused on symptomatic SVD. The combined effects of WMH and neuropsychiatric symptoms on cognitive and functional abilities are not known in the early-stages of SVD, where brain changes are detectible but no criteria are met for clinical diagnosis of stroke, dementia or significant disability (covert SVD).

We examined neuropsychiatric symptoms and domain-specific cognitive performance in subjects with varying degrees of WMH who were free from dementia and stroke, and were independent in their basic activities of daily activities. Specifically, we investigated whether a) WMH are associated with neuropsychiatric symptoms, b) neuropsychiatric symptoms are related to cognitive performance and instrumental activities of daily living (IADL), and c) neuropsychiatric symptoms moderate the relationship between WMH and cognitive and functional outcomes.

## METHODS

### Subjects and design

The Helsinki Small Vessel Disease Study is a cohort study investigating covert cerebral SVD through brain imaging, clinical and cognitive characteristics in older individuals. The study protocol has been described in detail previously.^14,15^ Briefly, the study included subjects who had recently undergone a brain scanning due to transient ischemic attack (26%), dizziness (18%), headache/migraine (10%), subjective cognitive complaints (4%), visual symptoms (11%), fall (7%), syncope (3%), or other reasons (20%). The subjects were recruited from the imaging registry of the Helsinki University Hospital in Finland between October 2016 and March 2020. A total of 152 subjects took part in the study including comprehensive neurological and neuropsychological assessments, self- and informant-filled questionnaires and a brain magnetic resonance imaging (MRI) with standard protocol carried out at three visits within approximately one month.

The inclusion criteria were: a) age 65-75 years at the time of enrolment; b) place of residence within the Helsinki and Uusimaa hospital district; c) occurrence of not more than minor, temporary and local neurological symptoms (having manifested 3 to 12 months before the enrolment), or no neurological symptoms at all; d) functional independence in daily activities as defined by a modified Ranking Scale score ^16^ of 0-2; and e) fluent Finnish language skills. The main exclusion criteria included significant neurological diseases, severe psychiatric disorders and factors that might hinder the administration of the neuropsychological tests (such as sight or hearing disabilities) or that might affect undergoing the MRI.

The study was approved by the Ethics Committee of the Helsinki University Hospital and conducted according to the Declaration of Helsinki. Informed written consent was received from each subject.

### Data availability statement

The data that support the findings of this study are available from the corresponding author upon reasonable request.

### Magnetic resonance imaging

A 3T MRI scanner with 32-channel head coil was used for the imaging. The protocol consisted of fast three plane localizer, 3D FLAIR SPACE, 3D T2 SPACE, 3D T1 MPRAGE, 3D gradient echo susceptibility weighted imaging sequence, 3D gradient echo sequence with magnetization transfer pulse on and off.

WMH of presumed vascular origin were defined on FLAIR sequences as hyperintense areas in the white matter without cavitation. A board certified neuroradiologist first evaluated WMH visually using the modified Fazekas scale ^17^ (0=none, 1=mild, 2=moderate and 3=severe), accounting for deep and subcortical WMH. Total WMH and grey matter (GM) volumes as well as periventricular and deep WMH volumes were segmented on FLAIR images with an automated multi-stage segmentation method based on the Expectation-Maximization algorithm.^18^ Three step method described earlier was used for the segmentation.^19^ Intracranial volume was used to normalise the total WMH and GM volume (ml) and the non-normality of the distribution was accounted for using a logarithmic transformation.

### Evaluation of cognitive functioning

Cognitive functioning was evaluated using a comprehensive neuropsychological assessment comprised of both paper-and-pencil and computerised tests described in detail previously.^15^ Briefly, the following domains were evaluated; processing speed, executive functions, working memory, memory and learning, visuospatial perception, and verbal reasoning. Domain scores were calculated by converting the raw scores (17 in total) into z scores and then the z scores were averaged within each domain. A global cognition score consisted of a standardised mean score comprised of the six domain scores. The scores used in the analyses of this study included global cognition, processing speed, executive functions and memory.

### Evaluation of neuropsychiatric symptoms

Neuropsychiatric symptoms were reported by a close informant Neuropsychiatric Inventory Questionnaire (NPI-Q).^20^ NPI-Q includes items on 12 neuropsychiatric symptoms (delusions, hallucinations, aggression, depression, anxiety, euphoria, apathy, disinhibition, irritability, motor disturbance, night time disruptive behaviours and appetite). For each symptom, the first question is whether the subject has the symptom (yes or no). NPI-Q total score is the sum of the occurrence of the 12 symptoms. A total of 134 informants filled in the NPI-Q.

### Evaluation of functional abilities

Instrumental activities of daily living were used as a measure of functional abilities. They were assessed with the short version of the Amsterdam Instrumental Activities of Daily Living (A-IADL) questionnaire also filled in by a subject’s close informant.^21^ The short version includes 30 questions assessing activities on areas such as taking care of household, work or finances, using appliances or a computer, as well as leisure activities. Scoring ranges from 0 (no difficulty) to 4 (unable to do activity) based on how performance is currently compared to the past performance. Item response theory is used to get a total score and higher scores show better functioning. A total of 132 informants filled in the A-IADL.

### Statistical analyses

First, we used Spearman’s correlations to examine the bivariate associations between NPI-Q total score and sex, age, years of educations as well as the MRI volumetrics (total, periventricular, deep WMH and GM volumes). Second, we ran a logistic regressions with the MRI volumetrics as the independent variables and the neuropsychiatric symptoms (present/absent) as the dependent variable. Third, we ran linear regression analyses with the neuropsychiatric symptoms (present/absent) as the independent variable and the cognitive or functional outcomes as the dependent variable (five analyses: global cognition, processing speed, executive functioning, memory and A-IADL). Fourth, we repeated the linear regression analyses by adding in the interaction terms (neuropsychiatric symptoms [present/absent] x WMH), as the independent variables and cognitive composite scores as well as A-IADL as the dependent variables. We then repeated the third and fourth steps for the most common neuropsychiatric symptoms (depression, irritability, night time disruptive behaviours, apathy and changes in appetite) as the independent variables. All analyses were controlled for sex, age and education (years). The false discovery rate correction was used for multiple analysis accounting for the five dependent variables in each set of analyses.

## RESULTS

### Subject characteristics

The study sample consisted of 134 subjects with informant filled NPI-Q, who did not differ from those with missing data (n=18) in sex, age, education, or WMH volume (all p-values>0.05). Exact number of subjects also varied depending on the NPI-Q item, most had 18 missing values, but night time disruptive behaviours had 21 missing values (usually due to the informant being unsure how to answer that particular question). Demographic information can be found in Table 1. Of the subjects, 62% had no neuropsychiatric symptoms, 16% had one symptom and 22% had two or more symptoms. The most common neuropsychiatric symptoms in our sample were depression (25%), irritability (21%), night time disruptive behaviours (15%), apathy (11%) and changes in appetite (11%). The occurrence of each neuropsychiatric symptom of the NPI-Q are detailed in Figure 1.

**Table 1.** Subject characteristics

|  | Subjects (n=152) |
| --- | --- |
| Demographics |  |
| Age (mean, sd) | 70.6 (2.9) |
| Sex (female/male, n) | 95/57 |
| Education (years, mean, sd) | 13.0 (4.5) |
| White matter hyperintensities |  |
| Fazekas score (none, mild, moderate, severe, n) | 14/76/42/20 |
| WMH volume (ml, mean, sd) | 10.5 (13.8) |

**Figure 1.**
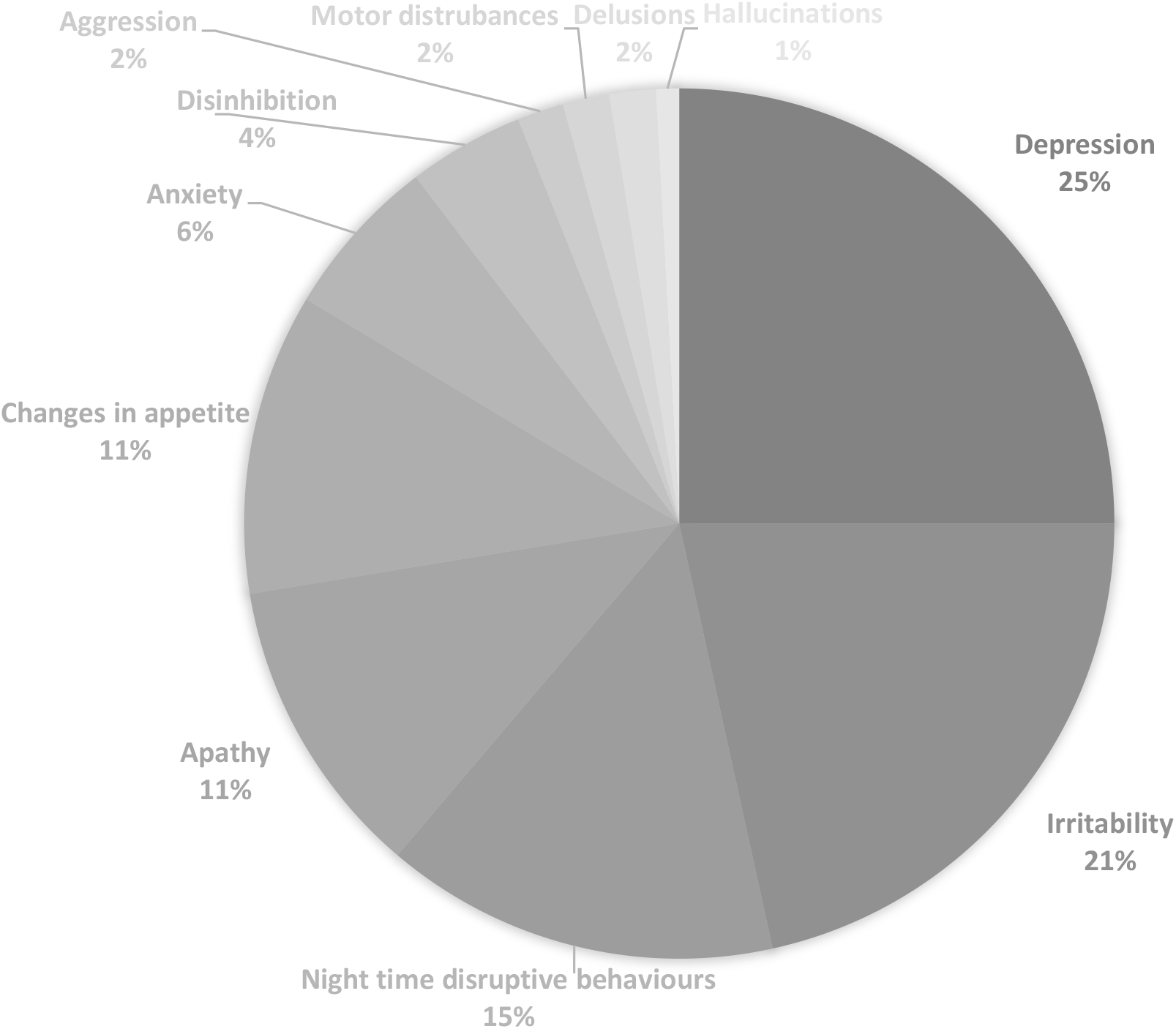
Occurrence of different neuropsychiatric symptoms (in percentages) based on the Neuropsychiatric Inventory Questionnaire (NPI-Q) from most common to least common clockwise.

### Relationships between NPI-Q and MRI volumetrics

NPI-Q total score correlated significantly with total WMH volume (r_s_=0.20, p=0.019), and more specifically with periventricular WMH volume (r_s_=0.20, p=0.024) and deep WMH volume (r_s_=0.19, p=0.028), but it was not related to total GM volume (r_s_=0.04), age, sex or years of education (all p>0.05). Total WMH volume was significantly associated with the presence of neuropsychiatric symptoms (binary variable) also after adjusting for age, sex and education (OR 2.89, CI 95% 1.13-7.43, p=0.027).

### Relationships between NPI-Q, cognitive scores and A-IADL

NPI-Q total score did not have significant bivariate correlations with cognitive composite scores (r_s_ values between −0.08 and −0.13, all p values>0.05). After adjusting for sex, age and education (years), there were no statistically significant associations between the presence of neuropsychiatric symptoms and cognitive functioning (standardised ß between −0.06 and - 0.13, all p-values >0.05). However, There were significant synergistic interactions between the presence of neuropsychiatric symptoms and WMH volume on all cognitive composite scores (Figure 2). Subjects with presence of any neuropsychiatric symptom together with higher levels of WMH volume had disproportionately low cognitive abilities.

**Figure 2.**
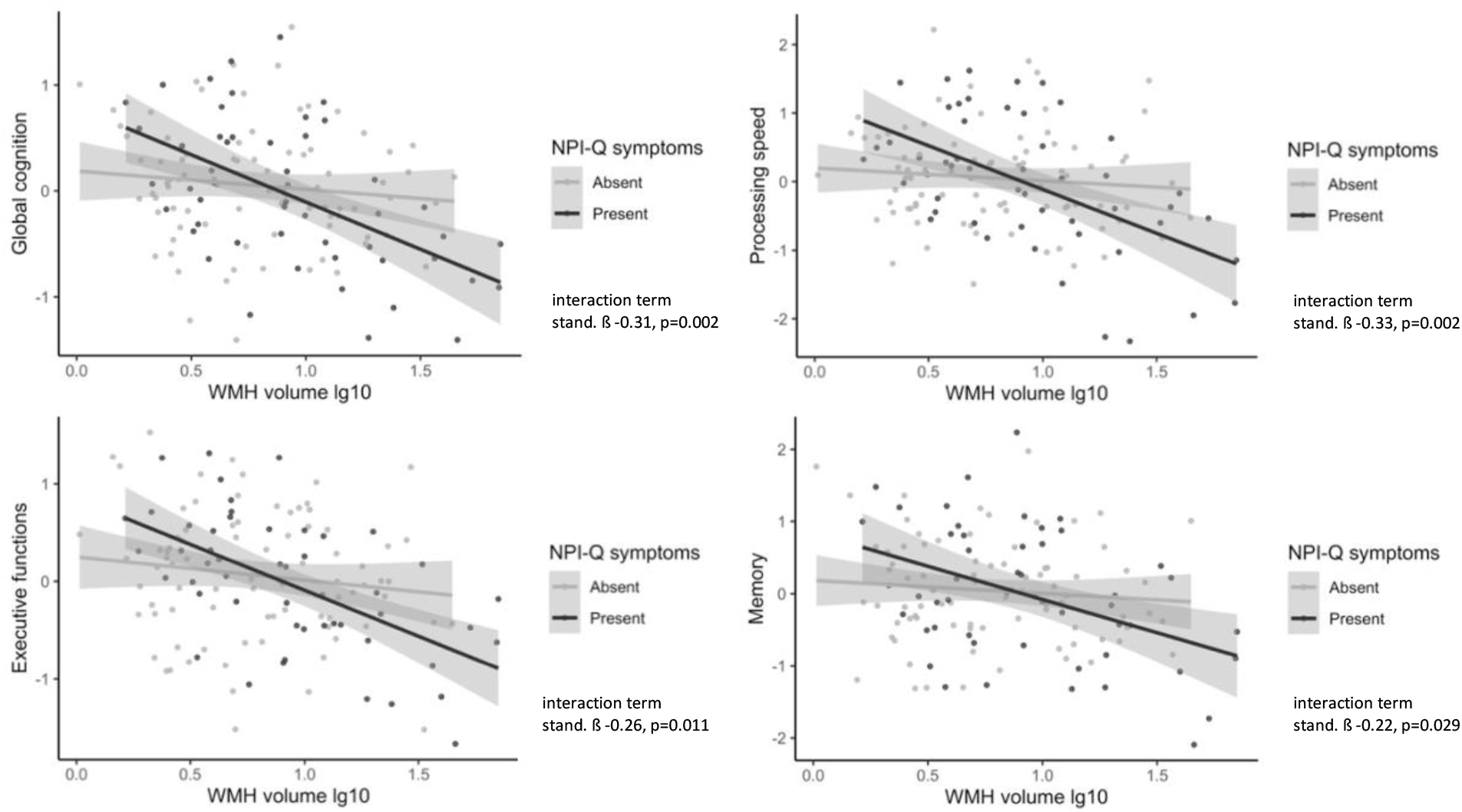
Interactions between white matter hyperintensities volume, presence or absence of informant evaluated neuropsychiatric symptoms based on the Neuropsychiatric Inventory Questionnaire (NPI-Q) and cognitive functioning. Grey areas represent 95% confidence intervals. All interactions were significant also after False Discovery Rate corrections.

NPI-Q total score correlated inversely with A-IADL score (r_s_=-0.41, p<0.001). After adjusting for sex, age and education, there was a significant association between the presence of neuropsychiatric symptoms and A-IADL (standardised ß −0.37, p<0.001). The interaction between the presence of neuropsychiatric symptoms and WMH volume on A-IADL was not significant (p-value>0.05).

### Associations between most common NPI-Q symptoms, cognitive scores and A-IADL

The presence of depression was significantly associated with global cognition (standardised ß −0.19, p<0.014) and A-IADL (standardised ß −0.43, p<0.001), but not with specific cognitive domains (standardised ß between −0.11 and −0.14, all p-values>0.05). There were significant interactions between the presence of depression and WMH volume on all the cognitive composite scores and A-IADL (Figures 3 and 4). Presence of depression together with higher levels of WMH volume associated with disproportionately poor cognitive composite and A-IADL scores. Irrespective of WMH, the presence of apathy was significantly associated with global cognition (standardised ß −0.29, p<0.001), processing speed (standardised ß −0.23, p=0.005), executive functions (standardised ß −0.19, p=0.017) and memory (standardised ß - 0.22, p=0.005) as well as A-IADL (standardised ß −0.46, p<0.001). The interactions between the presence of apathy, WMH and all the outcome variables were not significant (standardised ß between −0.07 and −0.19, all p-values>0.05). The associations between the other most common neuropsychiatric symptoms (irritability, night time disruptive behaviours and changes in appetite) and cognitive and functional outcomes were statistically non-significant (all p-values >0.05 after FDR correction).

**Figure 3.**
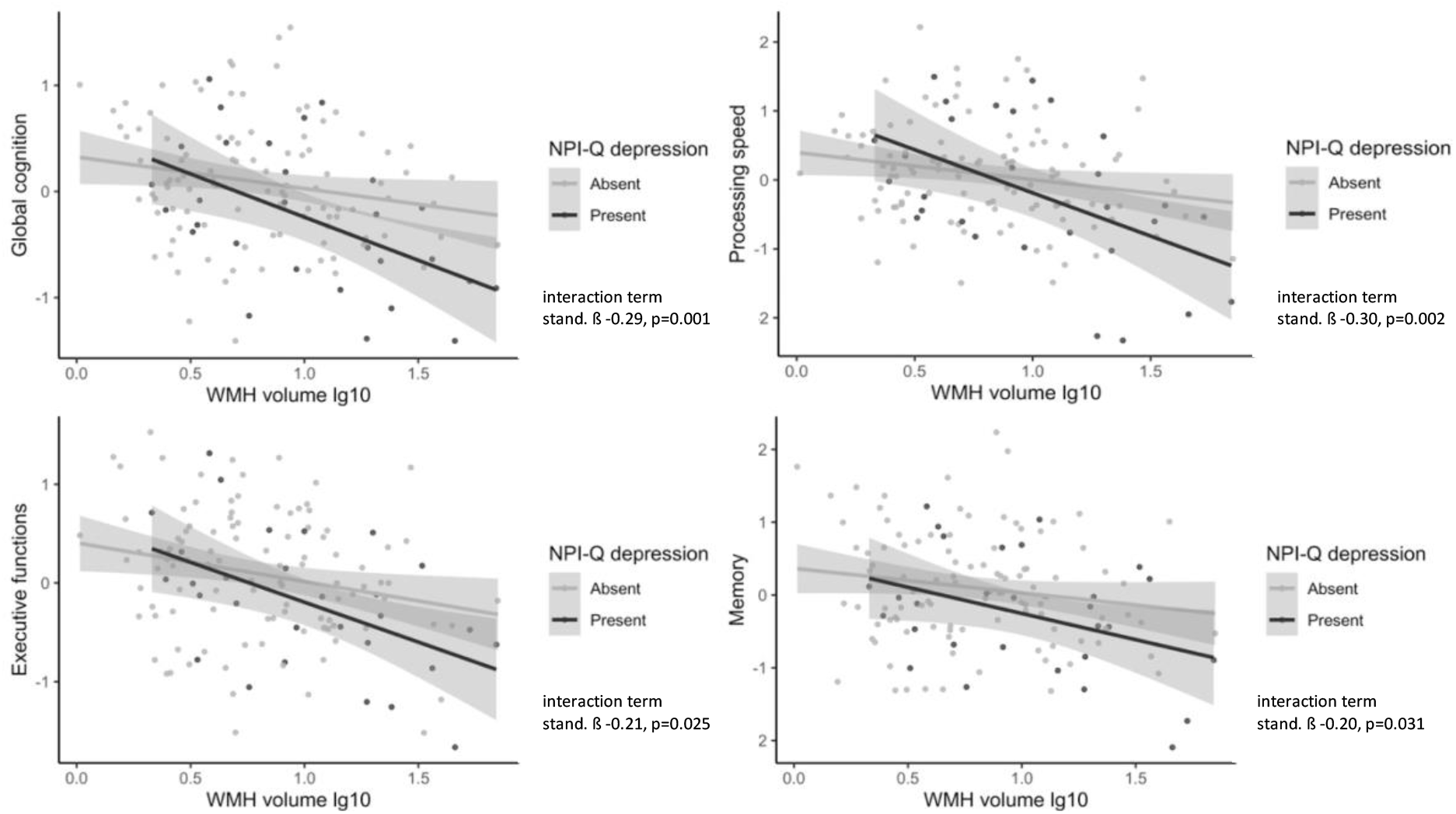
Interactions between white matter hyperintensity (WMH) volume and depressive symptoms group (based on the informant-evaluated Neuropsychiatric Inventory Questionnaire, NPI-Q) on cognitive functioning. Grey areas represent 95 % confidence intervals. All interactions were significant also after False Discovery Rate corrections.

**Figure 4.**
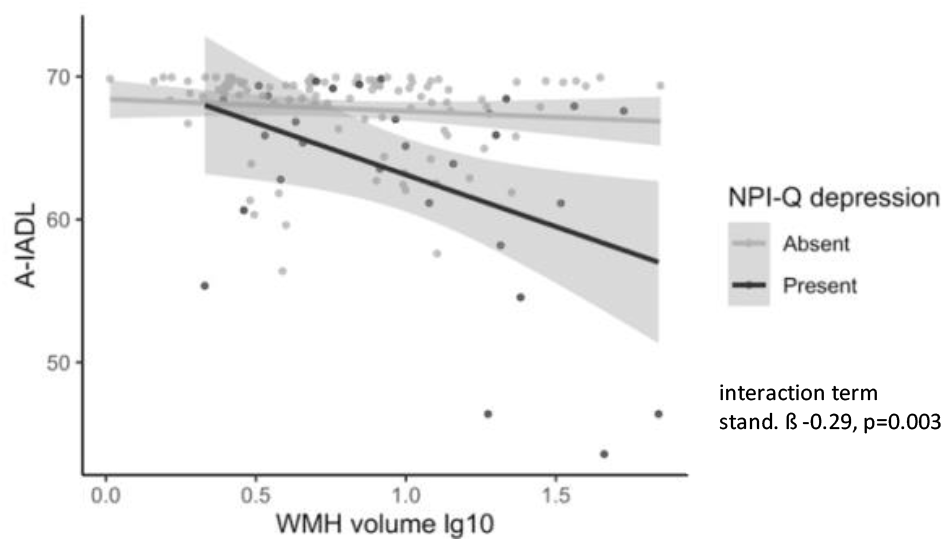
Interaction between white matter hyperintensity (WMH) volume and depressive symptoms group (based on the informant-evaluated Neuropsychiatric Inventory Questionnaire, NPI-Q) on functional abilities assessed with the Amsterdam Instrumental Activities of Daily Living questionnaire (A-IADL). Grey areas represent 95 % confidence intervals. Interaction was significant also after False Discovery Rate correction.

## DISCUSSION

We assessed the occurrence of neuropsychiatric symptoms and their relationship with MRI volumetrics and cognitive and functional abilities in covert SVD. The most common neuropsychiatric symptoms in our sample were depressive symptoms, irritability, night time disruptive behaviours, apathy and changes in appetite. Neuropsychiatric symptoms had significant correlations with total WMH volume, and with deep and periventricular WMH volume, but not with total GM volume. Neuropsychiatric symptoms were not directly related to cognitive functioning. Instead, the presence of neuropsychiatric symptoms together with higher total WMH volume was associated with exacerbated cognitive impairment in all the cognitive composite scores (global cognition, processing speed, executive functions and memory). Furthermore, neuropsychiatric symptoms were related to worse functional abilities.

Depressive symptoms were the most common symptoms in our sample. Depressive symptoms together with higher levels of WMH volumes were related to worse cognitive and functional abilities. Apathy symptoms were related to worse cognitive and functional performance irrespective of WMH severity. The other most common neuropsychiatric symptoms (irritability, night time disruptive behaviours and changes in appetite) were not related to cognitive or functional outcomes.

Previous meta-analyses have shown consistent associations between SVD brain pathology (WMH, lacunes, cerebral microbleeds and stroke) and depression, apathy, fatigue and delirium.^5,9^ In healthy individuals, there is tentative evidence with a small sample to suggest that WMH are related to significant preclinical neuropsychiatric symptoms.^23^ We also found evidence to suggest that WMH were related to the presence of neuropsychiatric symptoms already in the early stages of SVD without presence of clinical impairment. In our study, the overall occurrence of neuropsychiatric symptoms was 38%, which is a considerably high proportion considering the fact that the subjects represented cases with varying levels of WMH without stroke, dementia or disability.

Increasing neuropsychiatric symptoms have been previously shown to associate with SVD disease progression and cognitive decline over a two year period in a dementia-free elderly sample.^13^ Kan et al. found that at baseline, higher neuropsychiatric symptom incidence was associated with higher SVD-related brain pathology burden. The authors argued that neuropsychiatric symptoms could be used as a clinical marker of SVD progression. Our results were in line with these findings with significant associations between WMH volume and presence of neuropsychiatric symptoms. Furthermore, our results suggest that preclinical neuropsychiatric symptoms accelerate worse cognitive and functional impairment in covert SVD.

Previous research has identified differing results between apathy and depression in SVD. More precisely, apathy but not depression, has been shown to relate to white matter track connectivity and cognitive functioning. ^7,24^ Contrary to these previous results, we found depressive symptoms to be related to worse cognitive performance in the presence of higher levels of WMH, whereas apathy symptoms were related to worse cognitive outcomes irrespective of WMH. These previous studies used self-report questionnaires to determine the presence of apathy or depression. In our study, a close informant reported binary responses for each neuropsychiatric symptom. Neither of the evaluations represent a clinical diagnosis. We used solely the presence or absence of neuropsychiatric symptoms in our analyses and still found significant associations, highlighting the importance of informant interviews already at an early stage of SVD assessments in order to get a more thorough view on a patient’s ability to function.

A meta-analysis in mild cognitive impairment (MCI) has suggested that neuropsychiatric symptoms in the elderly might be an early sign of brain pathology and as such should be routinely screened.^25^ Apathy and irritability in particular have been linked to Alzheimer’s disease progression and cognitive decline. We found neuropsychiatric symptoms to be present already in covert SVD and have a significant role on subjects’ cognitive and functional abilities. Due to the potential to use neuropsychiatric symptoms as an early sign for future decline, more longitudinal research is needed.

To conclude, we found that neuropsychiatric symptoms are associated with WMH as the key brain changes of SVD and that they exacerbate the relationship between WMH and cognitive impairment already in the covert stages of the disease. Neuropsychiatric symptoms associate with worse functional abilities irrespective of WMH. Depressive symptoms in particular associate with worse cognitive and functional abilities, whereas apathy relates to worse functioning irrespective of WMH. Our results highlight the need to screen neuropsychiatric symptoms at an early stage of SVD assessment, as they are associated with worse cognitive and functional outcomes. Finally, our results highlight the importance of having an informant present at an evaluation, since even at the covert stage of SVD, an informant can aid in recognising those patients that are at a higher risk for worse outcomes.

## Acknowledgements

We thank Dr. Sietske S. A. Sikkes, Department of Clinical, Neuro and Developmental Psychology, VU University; Alzheimer Center Amsterdam, Department of Neurology, Amsterdam Neuroscience, Amsterdam UMC, Amsterdam, The Netherlands, for providing the A-IADL scale for the Helsinki Small Vessel Disease study.

The study was funded by the Helsinki and Uusimaa Hospital District, University of Helsinki and Päivikki and Sakari Sohlberg Foundation, Finland.

